# Estimation of time cost of anti-cancer drugs approved based on comparisons to best supportive care: a cross sectional analysis

**DOI:** 10.1101/2022.06.22.22276763

**Authors:** Vinay Prasad, Timothée Olivier, Emerson Y Chen, Alyson Haslam

## Abstract

**Background:** Financial costs from cancer treatment are increasingly recognized, but what has historically been underrecognized is the time cost of therapy. We sought to estimate the time burden of anti-cancer drugs approved based on comparisons to best supportive care (BCS), with the assumption that without this drug, a patient could have been treated with observation, home palliative care or hospice services, with minimal time seeking medical care.

**Methods:** We searched all FDA approvals (2009 - March 2022) for randomized trials that used BCS as a treatment option for an anti-tumor drug in the metastatic setting and abstracted data on treatment related activities. We then estimated time spent on these activities using previously calculated times.

**Results:** Of the 13 drugs tested against BSC, nine studies demonstrated an improvement in median OS (median 2.1 months). The median monthly time spent for patients in the intervention arm of BSC trials was 15.8 hours.

**Conclusion:** Time is a valuable resource for people who have cancer, but especially for patients who may have few to no remaining treatment options, and yet, we found that patients can spend up to 16 hours in anti-cancer drug related activities per month.

## Introduction

Cancer drugs may offer benefits, typically improvements in survival and quality of life, but also come with costs. Financial costs are increasingly recognized,^1^ but what has historically been underrecognized is the time cost of therapy. Patients receiving anti-cancer drugs spend time in transit to doctors’ visits, waiting for the provider, receiving infusions or drawing labs, and performing radiographic imaging, among other burdens. To date, there have been limited attempts to estimate the time cost of therapies, often focusing on single cancer histologies.^2,3^ We sought to estimate the time burden of anti-cancer drugs approved based on comparisons to best supportive care (BCS). This permitted us the assumption that without this drug, a patient could have been treated with observation, home palliative care or hospice services, with minimal time seeking medical care.

## Results

We found 13 approved drugs based on registration trials with BSC (with or without an active comparator). Eleven were blinded studies and used a placebo plus BSC control arm, and two studies were open-label and compared a drug to BSC only. The two open-label studies were in the maintenance setting (avelumab and pemetrexed). Five studies used an IV intervention, and the other eight were orally administered drugs.

Nine studies demonstrated an improvement in median OS (median 2.1 months; Table). Twelve studies demonstrated an improvement in PFS (median improvement of 1.6 months, with one study where median PFS was not reached).

**Table.**
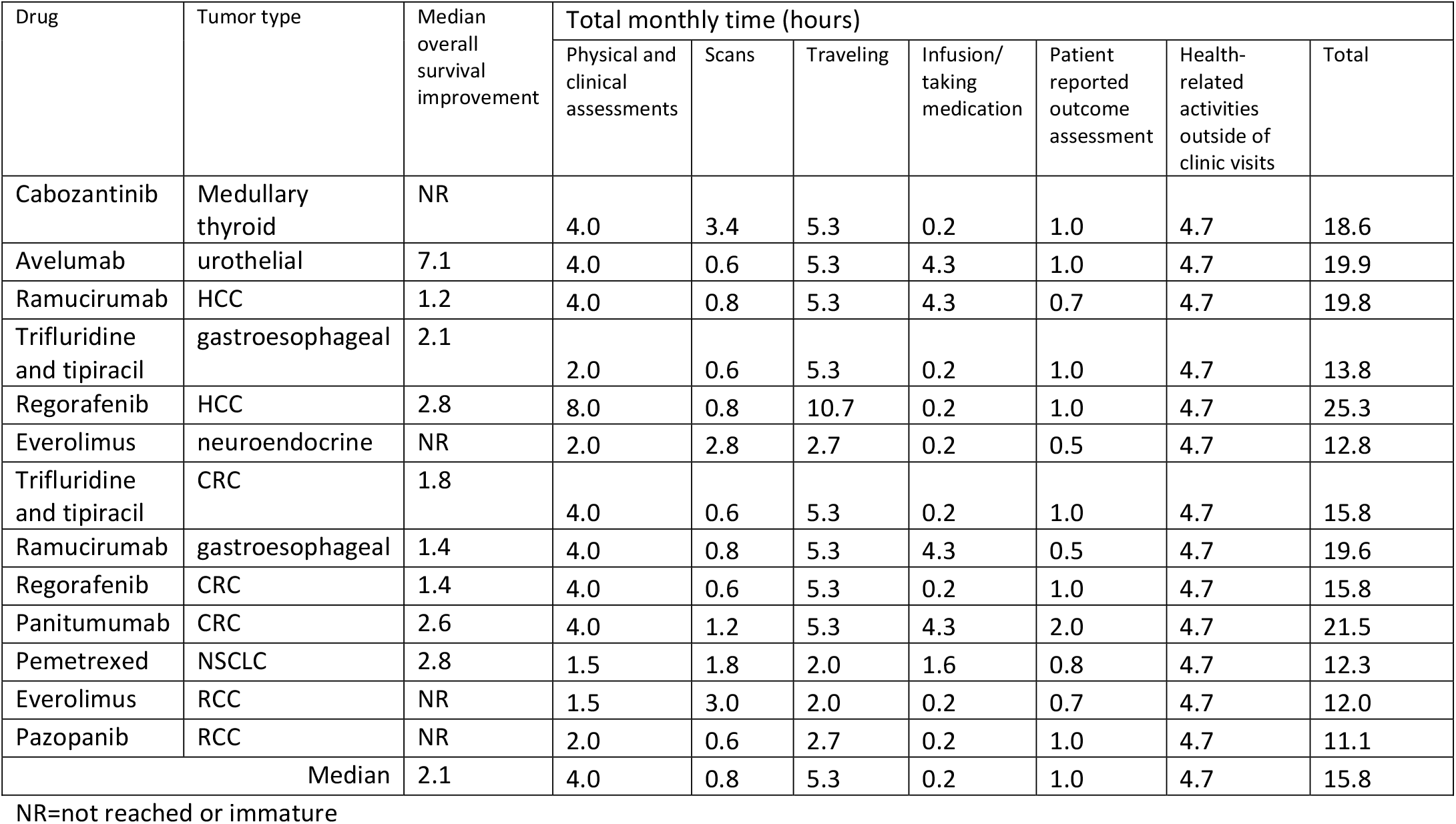
Time spent in health-related activities for drugs approved (2009-2022) and tested with best supportive care.

| Drug | Tumor type | Median overall survival improvement | Total monthly time (hours) |  |  |  |  |  |  |
| --- | --- | --- | --- | --- | --- | --- | --- | --- | --- |
|  |  |  | Physical and clinical assessments | Scans | Traveling | Infusion/ taking medication | Patient reported outcome assessment | Health-related activities outside of clinic visits | Total |
| Cabozantinib | Medullary thyroid | NR | 4.0 | 3.4 | 5.3 | 0.2 | 1.0 | 4.7 | 18.6 |
| Avelumab | urothelial | 7.1 | 4.0 | 0.6 | 5.3 | 4.3 | 1.0 | 4.7 | 19.9 |
| Ramucirumab | HCC | 1.2 | 4.0 | 0.8 | 5.3 | 4.3 | 0.7 | 4.7 | 19.8 |
| Trifluridine and tipiracil | gastroesophageal | 2.1 | 2.0 | 0.6 | 5.3 | 0.2 | 1.0 | 4.7 | 13.8 |
| Regorafenib | HCC | 2.8 | 8.0 | 0.8 | 10.7 | 0.2 | 1.0 | 4.7 | 25.3 |
| Everolimus | neuroendocrine | NR | 2.0 | 2.8 | 2.7 | 0.2 | 0.5 | 4.7 | 12.8 |
| Trifluridine and tipiracil | CRC | 1.8 | 4.0 | 0.6 | 5.3 | 0.2 | 1.0 | 4.7 | 15.8 |
| Ramucirumab | gastroesophageal | 1.4 | 4.0 | 0.8 | 5.3 | 4.3 | 0.5 | 4.7 | 19.6 |
| Regorafenib | CRC | 1.4 | 4.0 | 0.6 | 5.3 | 0.2 | 1.0 | 4.7 | 15.8 |
| Panitumumab | CRC | 2.6 | 4.0 | 1.2 | 5.3 | 4.3 | 2.0 | 4.7 | 21.5 |
| Pemetrexed | NSCLC | 2.8 | 1.5 | 1.8 | 2.0 | 1.6 | 0.8 | 4.7 | 12.3 |
| Everolimus | RCC | NR | 1.5 | 3.0 | 2.0 | 0.2 | 0.7 | 4.7 | 12.0 |
| Pazopanib | RCC | NR | 2.0 | 0.6 | 2.7 | 0.2 | 1.0 | 4.7 | 11.1 |
| Median |  | 2.1 | 4.0 | 0.8 | 5.3 | 0.2 | 1.0 | 4.7 | 15.8 |

NR=not reached or immature

The median monthly time spent for patients in the intervention arm of BSC trials was 15.8 hours (Figure). Traveling (5.3 hours/month) was the most time-intensive trial-related activities, followed by health-related activities outside of clinic time (4.7 hours/month), and physical/clinical assessments (4 hours/month). Time spent on treatment was the least time-intensive activity (median of 10 minutes/month) since most drugs were orally administered.

**Figure.**
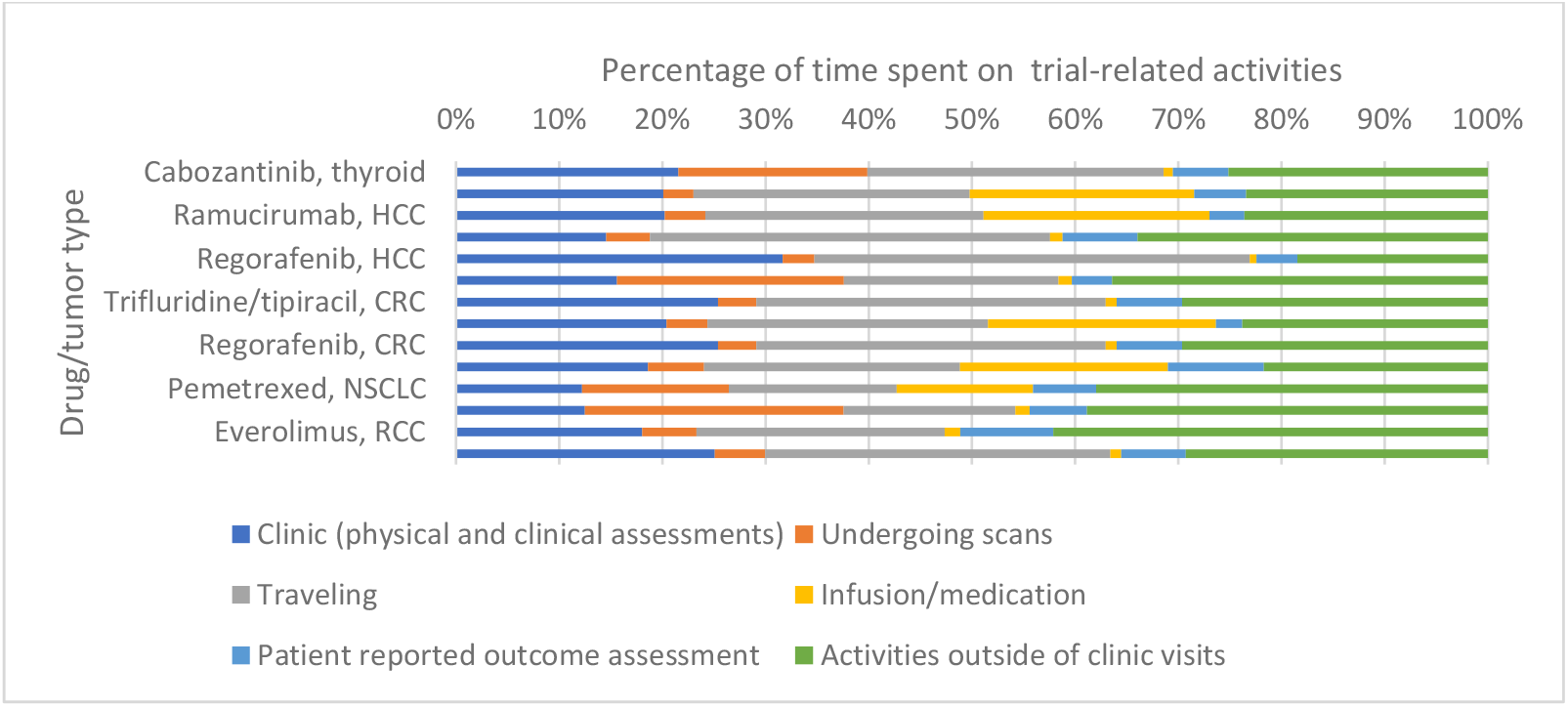
Percentage of time spent in health-related activities for drugs approved (2009-2022) and tested with best supportive care. HCC=hepatocellular carcinoma; CRC=colorectal cancer; NSCLC=non-small cell lung cancer; RCC=renal cell carcinoma.

## Discussion

Our estimates of almost 16 hours each month on treatment-related activities are likely underestimated due to inability to capture time spent being treated for adverse events.^4^ Additionally, these 16 hours may be distributed over at least 4-5 days per month, leading to fewer treatment-free ‘home days’.^5^ Time estimates were based on process mapping via direct observation in oncology clinics and patient surveys but applied to only the minimal frequency of visits for trial participation. Real-world data indicate that the frequency of clinic visits is more often than visits for trial assessments outlined in included trial protocols.^3^ One limitation is the few instances of BCS trials (N=13 trials), but this covered 13 years of FDA approvals.

Time is a valuable resource for people who have cancer, but especially for patients who may have few to no remaining treatment options. We selected trials where BSC was used, with or without an active comparator, which permitted us the assumption that without the drug, a patient could have spent minimal time seeking medical care. Indeed, we found that median improvement in OS was two months for patients on treatment. Patients spent almost 16 hours over a number of days in anti-cancer drug related activities per month. Future studies should prospectively assess actual time on care, including time for adverse events and hospitalizations for intervention and control arms, and calculate actual home days gained to further patients’ overall quality of life.

## Methods

In a cross-sectional analysis, we searched all FDA approvals (2009 - March 2022) for randomized trials that used BCS as a treatment option (control or intervention arm) for an anti-tumor drug in the metastatic setting. We abstracted study characteristics, including overall survival (OS) and progression-free survival (PFS) outcomes, open-label or not, intervention type (oral vs. IV), and type and frequency of assessments. We estimated time spent on trial-related activities (physical and clinical assessments, traveling, tumor assessments, treatment infusion, if applicable, and patient-reported assessments) for patients in the intervention trial arm by multiplying previously calculated times by the frequency of activity listed in the trial protocol.^3^

We conducted all analyses using R software, version 3.6.2 and Microsoft Excel. In accordance with 45 CFR §46.102(f), this study was not submitted for institutional review board approval because it involved publicly available data and did not involve individual patient data. We adhered to STROBE reporting criteria.

## Data Availability

All data generated or analyzed during this study are included in the manuscript and supporting file.

## Competing Interest

Vinay Prasad’s Disclosures; (research funding) Arnold Ventures; (royalties) Johns Hopkins Press, Medscape (honoraria) Grand Rounds/lectures from universities, medical centers, non-profits, professional societies, Youtube, and Substack (consulting); UnitedHealthcare; (speaking fees) Evicore; (other) Plenary Session podcast has Patreon backers. All other authors have no financial nor non-financial conflicts of interest to report.

## Data availability

All data for this are publicly available, and sources have been referenced in-text.

## Notes

### Funding Statement

This project was funded by Arnold Ventures

### Author Declarations

None required because we used publicly available data

